# Public attitudes toward sharing health data for artificial intelligence: Differences by data type and sector in the Health in Central Denmark cohort

**DOI:** 10.64898/2026.03.19.26348784

**Authors:** Jonas Rosborg Schaarup, Anders Aasted Isaksen, Adam Hulman

## Abstract

**Aims:** We aimed to examine public perceptions of sharing various types of health data relevant for AI development, including electronic health records, audio recordings of consultations, medical images, and genetic information, with actors from either the public or the private sectors.

**Methods:** We analysed data from 38,740 participants of the Health in Central Denmark survey conducted in 2024. Participants were asked whether they would share different types of health data with an AI solution in healthcare. Each participant was randomised to either of two versions of the scenario and question where the AI application was developed in the public or private sector. Descriptive results (proportions and percentages) were weighted to represent the background population of approx. 1 million people in the Central Denmark Region. The association between randomization group (data recipient) and data sharing attitude (“Yes”, “No”, “Don’t know”) was analysed using multinomial logistic regression with “Don’t know” as reference category.

**Results:** Participants were most willing to share medical images (46%), followed by text from patient journals (39%), genetic information (35%), and audio recordings (27%). There were 12-16% higher proportions of willingness to share with public institutions than with private institutions. A high level of uncertainty was observed for all data types (29-36%) regardless of data recipient. Odds ratios ranged from 1.37 to 1.78 for responding “Yes”, and from 0.51 to 0.67 for responding “No” to sharing data with public institutions compared to private institutions.

**Conclusions:** Public acceptance of health data sharing for AI depends on both the perceived sensitivity of the data and the institutional context of use. Strong public governance, transparent safeguards, and clear communication about data use may be important for maintaining trust and enabling responsible development of AI in healthcare.

## Introduction

Vast amounts of data are routinely collected within healthcare systems and are increasingly used to support clinical decision-making, quality assurance, and research [1, 2]. Recent advances in artificial intelligence (AI) have expanded the potential applications of these data, enabling the analysis of electronic health records with large language models (LLMs), transcription and summarization of audio recordings from clinical consultations, automated detection of abnormalities in medical images, and integration of genetic information to personalize care, among other applications [3]. However, the development of clinically relevant AI-based applications depends on access to large, high-quality datasets that reflect real-world patient populations and care pathways. Facilitating such access is therefore crucial to support innovation that may ultimately improve patient outcomes and enable more efficient allocation of healthcare resources.

Public support for data sharing in health research is generally high, especially in Nordic countries, although willingness varies depending on contextual factors such as data type, perceived sensitivity, purpose of use, and the identity of the data recipient [4]. Prior studies have consistently shown higher levels of trust in public sector institutions compared with private or commercial actors, suggesting that the perceived motivation of data users (e.g. public benefit versus commercial interest) plays a key role in shaping attitudes [5]. The type of data also influences willingness to share. Genetic data is often regarded as more sensitive than administrative or clinical records. A large international study on genetic data donation found that familiarity with genetics was associated with a higher willingness to share DNA data for research, highlighting the importance of public understanding and literacy in this domain [6]. However, it is an open question whether people’s willingness would differ if such data were used for AI applications.

Ethical and governance considerations are central to public acceptance of secondary health data use. AI technologies introduce additional challenges that go beyond traditional research ethics. Concerns include privacy and data protection [7], potential bias and discrimination in algorithmic outputs [8], lack of explainability (“black box” models), and uncertainty regarding accountability when AI systems make or influence clinical decisions [9]. Moreover, general public understanding of AI remains limited, and many still perceive it as a complex or even mysterious technology [10]. This limited familiarity may increase perceptions of risk and uncertainty, reducing trust in AI-driven health data use compared with more conventional research methods.

In Denmark, trust in public institutions is generally high, and citizens tend to express relatively low concern about data use for research purposes [4]. However, recent public debates and media coverage of data-related controversies have highlighted tensions between innovation and privacy, underscoring the need for continuous attention to public perceptions [11, 12].

Previous studies on data sharing preferences have rarely focused explicitly on AI applications or compared attitudes across different data types and data recipients. The present study addresses this gap by examining public perceptions of sharing various types of health data relevant for AI development, including electronic health records, audio recordings of consultations, medical images, and genetic information, with actors from both the public and private sectors.

## Methods

### Data sources

#### Survey data

Health in Central Denmark (HICD) is a digital and postal questionnaire survey initially conducted in 2020 on all inhabitants of Central Denmark Region aged 18 to 74 years identified with diabetes in register data on 31 December 2018, and an equally-sized group of people without diabetes (matched by sex, age, and municipality) [13].

In 2024, a new survey was conducted, inviting 136,229 participants aged 18 to 92 years, providing the basis of our current study. As part of the HICD survey in 2024, we collected data on willingness to share different modalities of healthcare data for use with artificial intelligence applications across the public and private sectors. For the question whether participants felt comfortable about an AI computer program analysing their data, participants were randomised to two versions of the survey question. The only difference between the two versions was whether the developer and data recipient was a public vs. private organisation. All participants were then asked to respond to four items representing different types of data: 1) electronic health records (text and tabular data), 2) medical images (scans), 3) audio recordings of consultations, and 4) genetic information. Participants were presented with three possible responses to express their data sharing preferences: “Yes”, “No”, “Don’t know”. The original survey and its English translation are available elsewhere [14].

HICD was approved by Statistics Denmark and the Danish Health Data Authority and registered in the Central Denmark Region internal register of research projects (reg. no. 1-16-02-165-20). Informed consent was collected from participants. Observational studies based on questionnaire and register data are exempt from ethical approval in Denmark.

#### Register-based information

We combined the survey data with administrative and healthcare data from nationwide Danish registers. Information on age, sex, and education level (defined by the total duration of education attained) was obtained from the Danish Civil Registration System and the Danish Population Education Register. Diabetes status was defined as prevalent “Type 1 diabetes” (T1D), “Type 2 diabetes” (T2D), or “No diabetes” at baseline based on hemoglobin A1c measurements ≥48 mmol/mol (6.5%), hospital diagnoses of diabetes, diabetes-specific podiatrist services, and purchases of glucose-lowering drugs [15]. These measures were derived for the entire background population corresponding to the survey sample, including all residents aged 18-92 years in the Central Denmark Region as of 1 January 2024.

### Statistical analysis

Participants who responded to all questions regarding types of data sharing were included in the analyses. To account for the survey’s sampling design and differential survey participation by demographic and clinical characteristics, we estimated each individual’s probability of survey response in a logistic regression model using age, sex, education level, and diabetes status as independent variables on all 1,010,111 individuals in the background population. These probabilities were used to compute stabilized weights for each respondent, truncated at the 95th percentile to reduce the influence of extreme values. These weights were applied in all analyses, and weighted results were presented throughout the main text, unless stated otherwise. Unweighted descriptive statistics and estimates are shown in the supplementary materials.

The distribution of study population characteristics (age, sex, education level, diabetes status) and data sharing attitudes is presented overall and tabulated as frequencies (%) or mean (SD). Univariable multinomial logistic regression was used to model the association between randomisation group (public vs. private sector) as data recipient and the attitude toward sharing healthcare data for use in clinical AI (with “Don’t know” as the reference outcome).

## Results

### Study population characteristics

From the survey population (n=136,229), 39,048 (29%) respondents completed the questionnaire about data sharing. Register-based information on education level was missing on 308 individiduals, who were excluded from analyses (Figure 1). The mean (SD) age was 56 (13) years, and 53% were women. A full description of demographic details by randomization to public and private sector scenarios is shown in Table 1. No systematic differences were observed between the randomization groups.

**Table 1.**
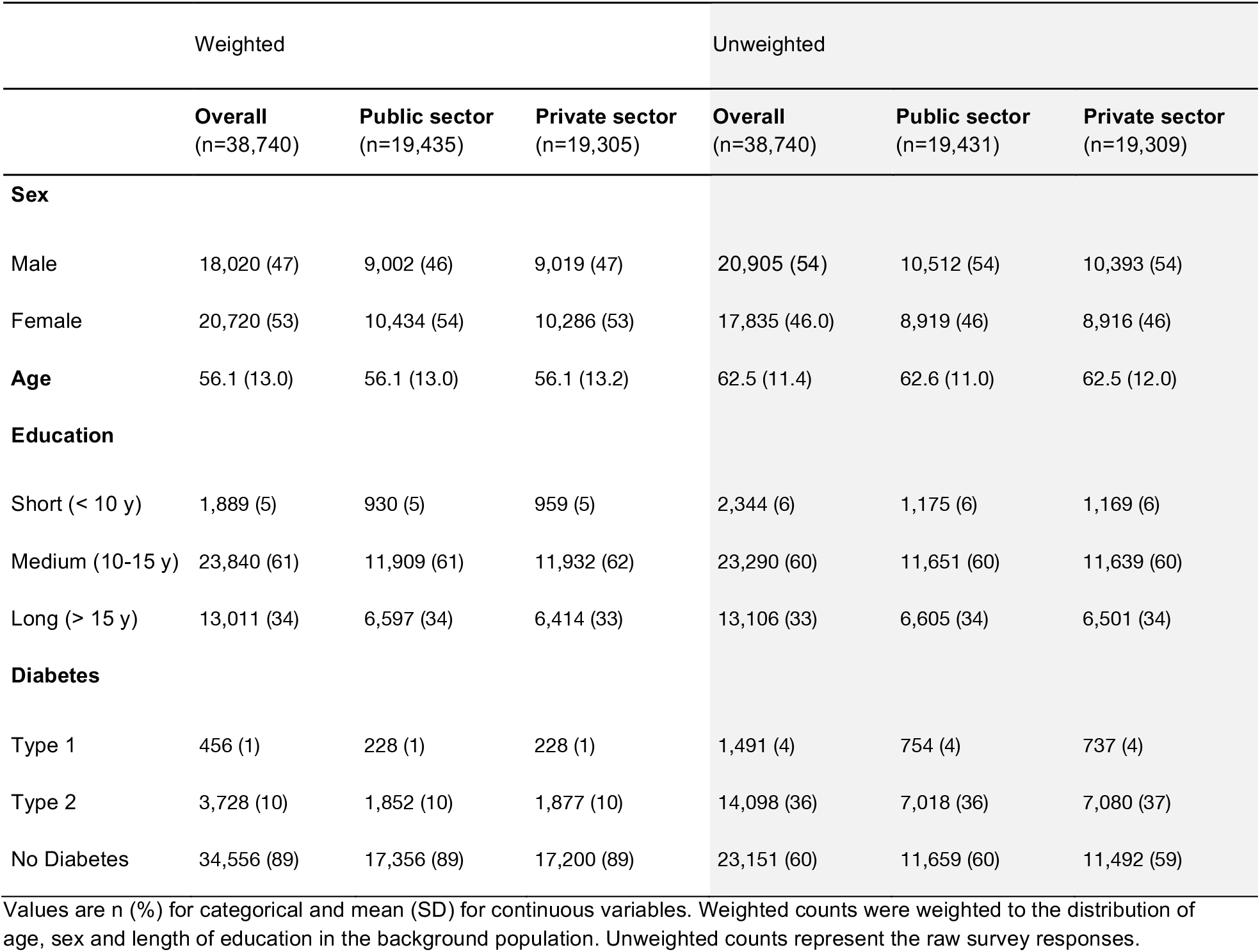
Weighted and unweighted population characteristics by randomization group.

**Figure 1.**
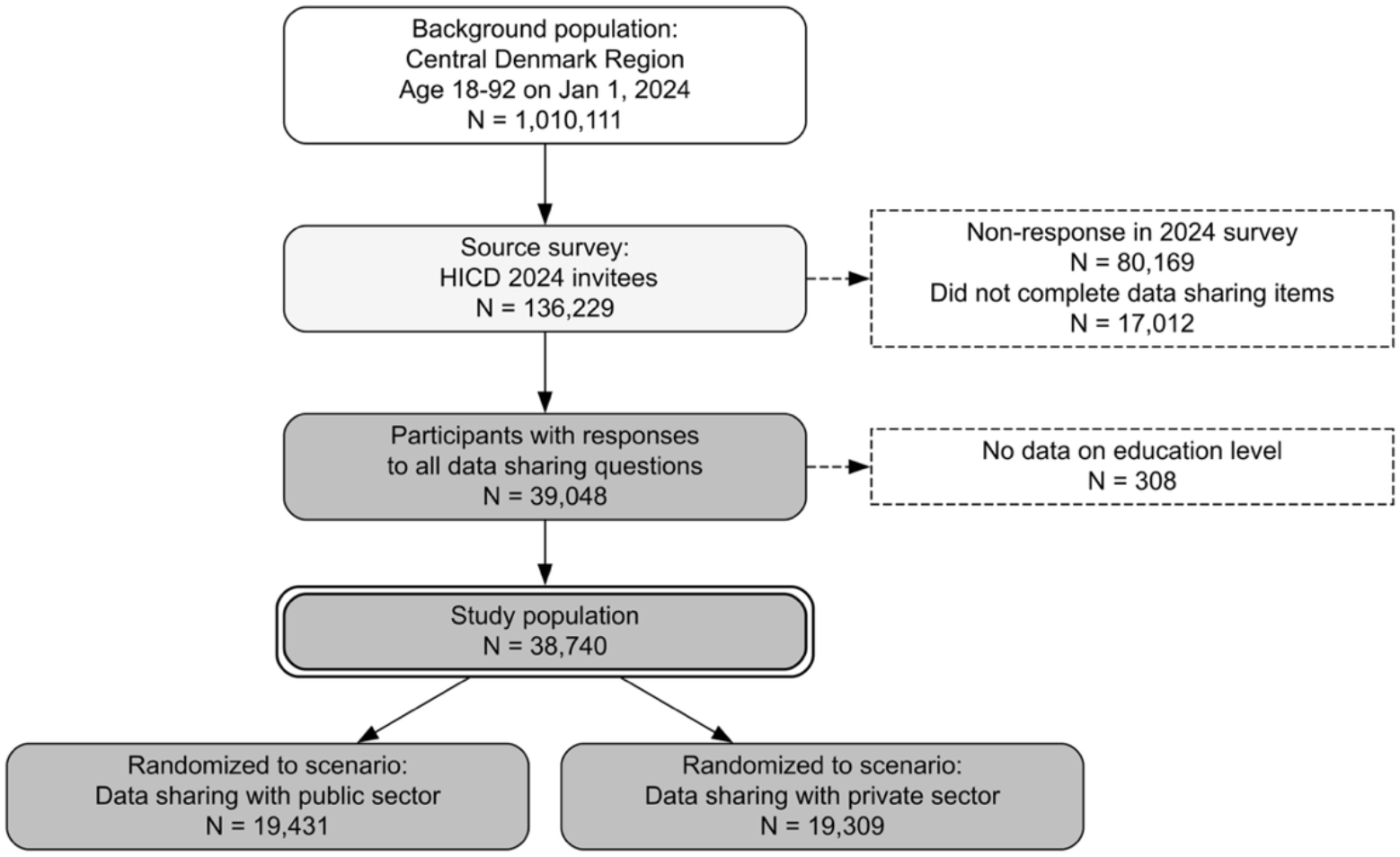
Flowchart and randomization by questions

### Data sharing attitudes

Overall, the types of data respondents were most willing to share were medical images (46%), followed by text from patient journals (39%), genetic information (35%), and audio recordings (27%). These figures varied between public and private sectors, with a 12-16% higher proportion of willingness to share with public institutions than with private institutions. A high level of uncertainty was observed in the sharing of genetic information (36%) and audio recordings (34%). No difference in magnitude (1-2%) was observed in the ‘Don’t know’ category between public and private sectors across all data types. Full description of data sharing attitudes by public and private sectors are shown in Table 2 (unweighted estimates are presented in Table S1). With “Don’t know” as the reference category, the odds ratios ranged from 1.37 to 1.78 for responding “Yes”, and from 0.51 to 0.67 for responding “No” to sharing data with public institutions compared to private institutions (Table 3). Unweighted estimates are presented in Table S2.

**Table 2.**
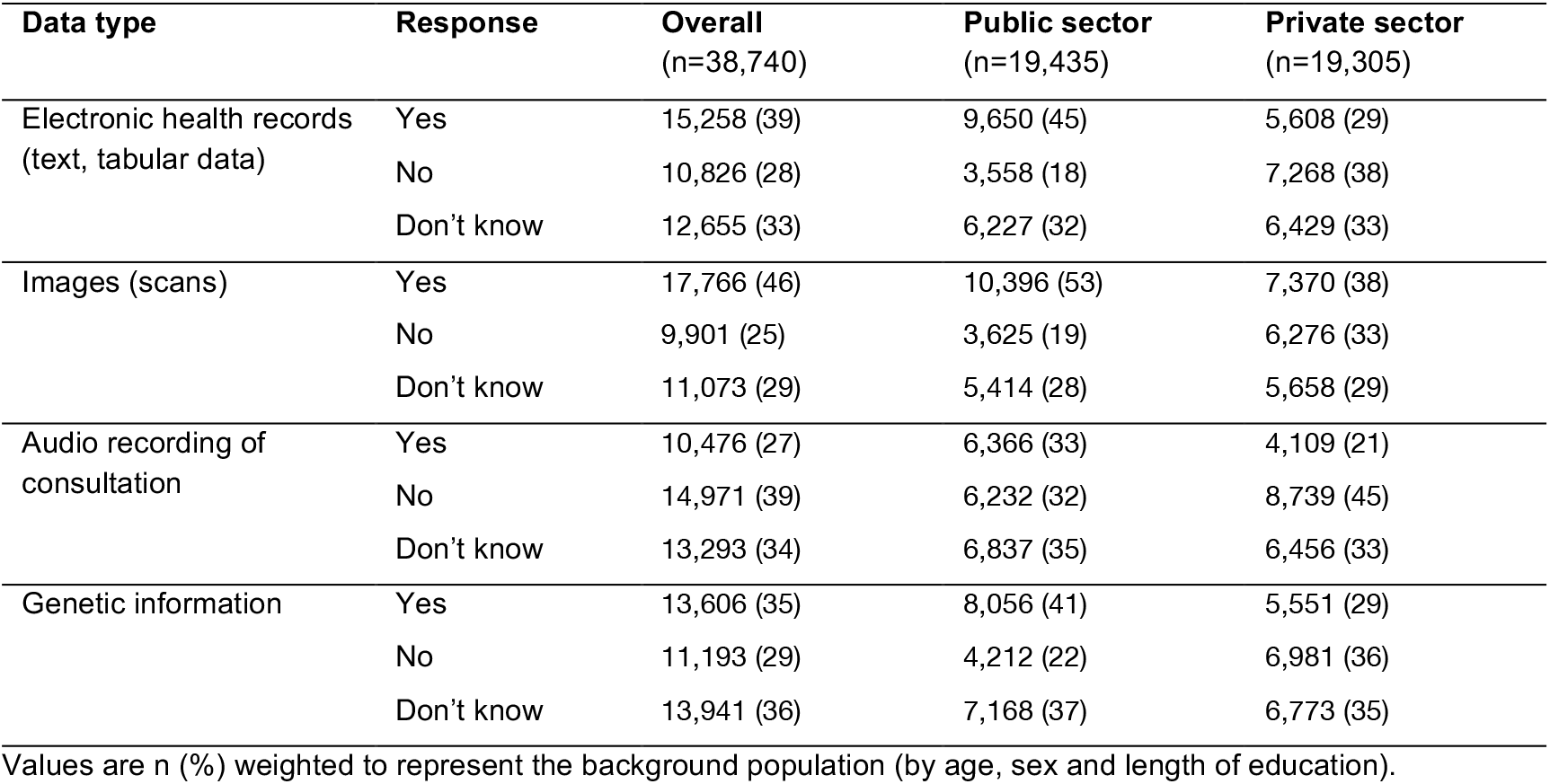
Distribution of data sharing attitudes by data type and sector.

| Data type | Response | Overall<br>(n=38,740) | Public sector<br>(n=19,435) | Private sector<br>(n=19,305) |
| --- | --- | --- | --- | --- |
| Electronic health records<br>(text, tabular data) | Yes | 15,258 (39) | 9,650 (45) | 5,608 (29) |
|  | No | 10,826 (28) | 3,558 (18) | 7,268 (38) |
|  | Don't know | 12,655 (33) | 6,227 (32) | 6,429 (33) |
| Images (scans) | Yes | 17,766 (46) | 10,396 (53) | 7,370 (38) |
|  | No | 9,901 (25) | 3,625 (19) | 6,276 (33) |
|  | Don't know | 11,073 (29) | 5,414 (28) | 5,658 (29) |
| Audio recording of<br>consultation | Yes | 10,476 (27) | 6,366 (33) | 4,109 (21) |
|  | No | 14,971 (39) | 6,232 (32) | 8,739 (45) |
|  | Don't know | 13,293 (34) | 6,837 (35) | 6,456 (33) |
| Genetic information | Yes | 13,606 (35) | 8,056 (41) | 5,551 (29) |
|  | No | 11,193 (29) | 4,212 (22) | 6,981 (36) |
|  | Don't know | 13,941 (36) | 7,168 (37) | 6,773 (35) |

Values are n (%) weighted to represent the background population (by age, sex and length of education).

**Table 3.**
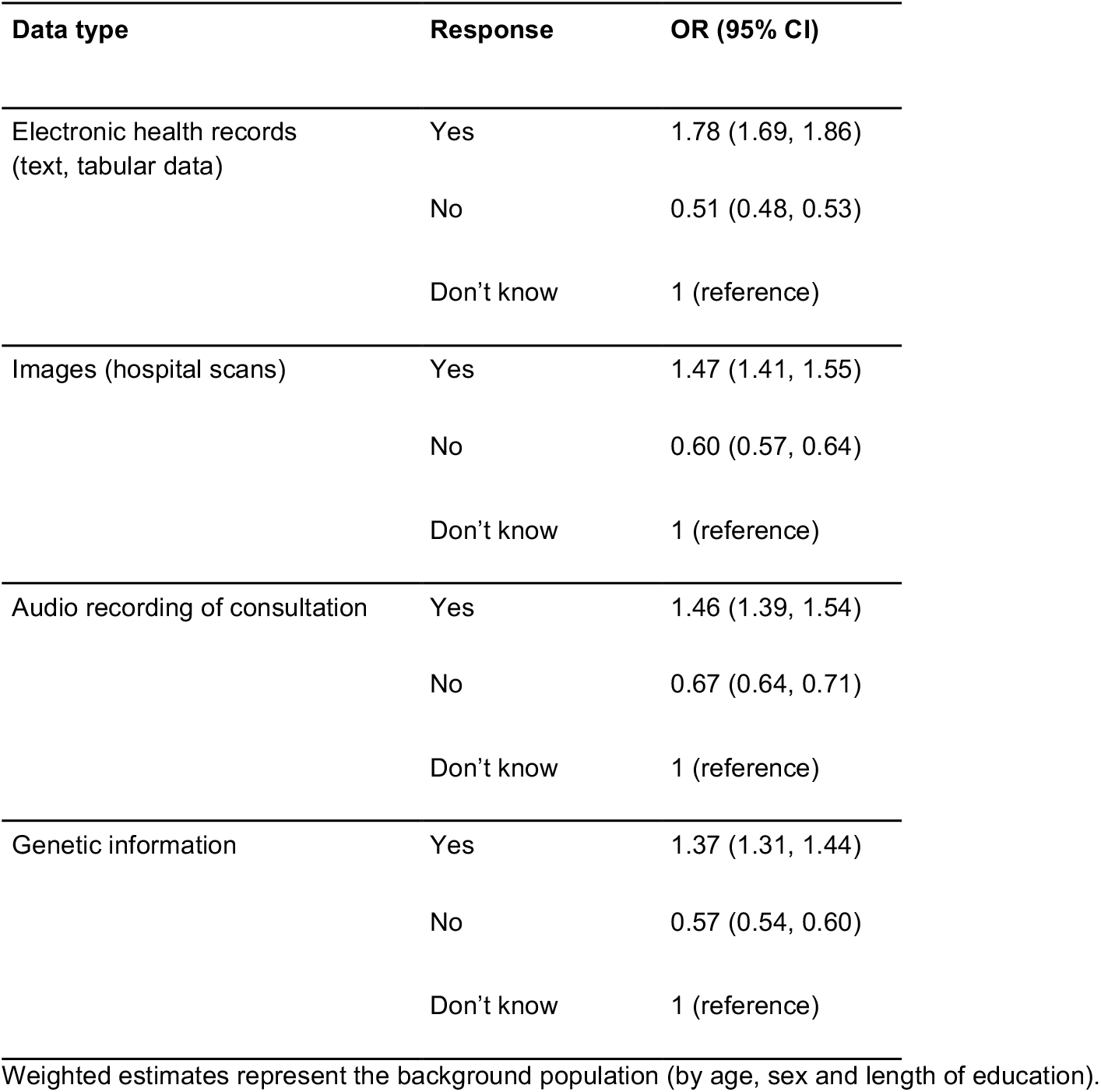
Association between data recipient sector (public vs. private [reference]) and willingness to share data.

| Data type | Response | OR (95% CI) |
| --- | --- | --- |
| Electronic health records<br>(text, tabular data) | Yes | 1.78 (1.69, 1.86) |
|  | No | 0.51 (0.48, 0.53) |
|  | Don't know | 1 (reference) |
| Images (hospital scans) | Yes | 1.47 (1.41, 1.55) |
|  | No | 0.60 (0.57, 0.64) |
|  | Don't know | 1 (reference) |
| Audio recording of consultation | Yes | 1.46 (1.39, 1.54) |
|  | No | 0.67 (0.64, 0.71) |
|  | Don't know | 1 (reference) |
| Genetic information | Yes | 1.37 (1.31, 1.44) |
|  | No | 0.57 (0.54, 0.60) |
|  | Don't know | 1 (reference) |

Weighted estimates represent the background population (by age, sex and length of education).

## Discussion

The development and success of AI solutions in healthcare depend on public trust and willingness to share personal health information. In one of the largest population-based surveys, and one of the first to focus on data sharing specifically for use in AI, we found that willingness to share data varied substantially by both data type and data recipient. Overall, respondents were most comfortable sharing medical images and electronic health record text, whereas genetic information and audio recordings of consultations elicited greater reluctance and uncertainty. Across all modalities, willingness to share was consistently higher when AI systems were developed and operated within the public healthcare sector compared with private organisations, with 12-16% greater acceptance and lower odds of refusal.

In this study, public willingness to share healthcare data for use in AI was much lower than figures presented in a systematic review on willingness to share health data for secondary purposes, which reported a pooled estimate of 77% from 65 studies across many countries [5]. The review was not limited to data sharing for use in AI, and skepticism in the context of AI may contribute to the lower willingness to share data found in our study. This hypothesis aligns with results from a survey of 12,153 patients in Canada [16], where 63% reported privacy concerns regarding AI use in healthcare, although the levels of outright unwillingness to share data for use in AI were not as high in our study (between 25% to 39% depending on data type).

### Data type and setting

Despite showing the highest level of willingness among the four data modalities included in the survey, willingness to share medical images was lower in our study (53% for sharing with public sector) compared to a study from Canada among patients subjected to ultrasound, computed tomography, or magnetic resonance imaging, 76% of which were willing to share data for research purposes [17, 18]. This difference in willingness to share imaging data may reflect the different contexts of the two surveys: ours being conducted electronically in the general public, whereas the other involved physically contacting patients at the clinic where they underwent medical imaging. The close relation between participants and the imaging data processor may have positively influenced the level of trust.

In our study, the proportion of individuals willing to share genetic data with stakeholders from the public and private sector was lower (41% and 29%, respectively) compared to a large survey across 22 countries on willingness to share genetic data, which reported figures of 49% and 32% for sharing with non-profit and for-profit researchers, respectively. This difference appeared to be due to a higher level of uncertainty in our study, as the proportion of individuals expressing unwillingness to share data was similar between our study and the review [6].

Roughly one-third of participants were against sharing audio from consultations for use in AI, a level much lower than in a Canadian survey, where 62% were against using AI scribes in their consultations [19]. In contrast, a study from California found a much lower level of concern, as only 15% reported privacy concerns [20]. That study was conducted among patients in a healthcare center explicitly to learn their preferences and inform a planned implementation of AI scribes in the center, and this explicit user involvement may have had a positive influence on patients’ perception.

The prevalence of uncertainty about sharing data for use in AI was high across all data types (roughly one-third of respondents). This figure was higher compared to a previous study from the UK, in which 18% express uncertainty about their willingness to transfer anonymized health data for AI research purposes [21]. The high level of uncertainty in our study may suggest that a large part of the Danish public experience either a lack of engagement on the issue or a lack of knowledge to make an informed opinion on the matter. The latter interpretation is supported by previous research showing that higher digital literacy is associated with a higher willingness to share health data for secondary use [22]. In a previous study conducted in our survey population, more than 90% of respondents were aware of AI and three-quarters had heard of ChatGPT, but only one-in-five had personal experience using AI tools such as ChatGPT [23]. Consequently, the “black box” nature of AI may amplify the perceived complexity and risk of sharing sensitive data types. However, emerging evidence suggests that this uncertainty can be impacted by exposure to AI technology, as direct experience with generative AI tools, such as ChatGPT, has been associated with decreased uncertainty regarding the risks and benefits of AI in healthcare [24]. This implies that as public familiarity and digital literacy regarding AI tools increase, the currently high levels of uncertainty may diminish, but it is uncertain whether this will translate into higher willingness or opposition to sharing data. A qualitative study from the UK discussed the value of large research databases. The results suggested that people prefer clear and concise information about the purpose of AI research, how data are anonymised and stored, who will access the data, and the conditions under which data may be shared [25]. The uncertainty observed in our study therefore underscores the need for ongoing public engagement and transparent communication about these aspects. As newer data modalities are introduced, such engagement will be essential to define consent and to maintain societal trust in health data research.

### Data recipient

Our study demonstrated greater willingness to share health data with public institutions than with private organisations regardless of data type, extending previous findings on data sharing attitudes in healthcare and health research to the context of AI applications. This pattern aligns with international evidence showing higher levels of trust in public or non-profit actors compared with commercial entities [5, 26]. For instance, a comparable survey from the UK found a dramatic divide in willingness to share health data for AI, reporting 66–78% acceptance for public institutions versus only 26% for commercial organisations [21]. Notably, that study found uncertainty to be largely concentrated around the private sector (34% vs 14–21% for public), whereas our results indicate a consistently high level of uncertainty across both sectors. This suggests that while the preference for public governance is universal, our Danish cohort exhibits a high level of uncertainty regarding health data sharing for AI, irrespective of the data recipient.

In Denmark, concerns about commercial motives appear particularly salient: in a previous national survey [4], 63% of respondents agreed that they were worried financial profit might influence how commercial companies use health data, representing the most frequently cited concern compared with issues such as data security, cross-border transfer, or discrimination. Together, these findings suggest that perceptions of institutional intent and public benefit are central determinants of acceptability in line with public priorities seen qualitative interviews in the UK [25].

At the same time, much of the current AI innovation landscape is driven by the private sector. More than a thousand AI-based software-as-medical-device tools have been approved by the FDA, the majority by for-profit companies, and a substantial share of AI research and infrastructure is led by large technology firms. This creates a structural tension: while health data are largely generated and governed within public healthcare systems, many of the technical capabilities required to develop and deploy AI solutions reside in private industry. Bridging this divide requires collaboration models that leverage complementary strengths without undermining public trust. Clear governance frameworks are therefore essential, including transparent agreements on data access and ownership, strict purpose limitation, secure analysis environments, and safeguards to prevent secondary or non-health-related uses.

European regulatory initiatives provide an important foundation for such approaches. The General Data Protection Regulation (GDPR) establishes strong protections for personal data and sets conditions for lawful processing, including health information, which may constrain certain uses of externally hosted or proprietary AI systems [27]. Building on this, the European Health Data Space seeks to enable secure secondary use of health data for research and innovation through controlled access mechanisms, anonymization or pseudonymization, and processing within secure environments [28]. In parallel, the European AI Act introduces risk-based requirements for AI systems, with healthcare applications classified as high risk and therefore subject to enhanced obligations regarding transparency, documentation, human oversight, and accountability [29]. Together, these frameworks illustrate an effort to balance innovation with individual rights and societal expectations. Our findings underscore the importance of such safeguards, suggesting that robust public governance and clear accountability mechanisms are not only legal necessities but also key enablers of public acceptance of AI-driven health data use.

### Strengths and limitations

Our study included almost 40,000 participants, which is the largest number according to a recent systematic review on health data sharing attitudes [5]. This large sample size allowed us to implement a strong design and randomise participants to different versions of the same question and thus avoid potential confounding when investigating the effect of who the data recipient was. We also minimised the effect of selection bias by reporting weighted estimates based on register-based information from almost one million people in the background population. We have incorporated emerging data types in our survey, e.g. audio recordings during consultations, that are receiving more attention with recent developments in AI. On the other hand, we must acknowledge that participants were presented hypothetical scenarios in an online survey, which might influence their attitudes. These scenarios did not specify the clinical context or the precise purpose of data use, both of which are recognised as substantial factors in shaping public trust [25]. We hypothesise that participants would be more willing to share data in a real-world scenario with purpose-specific cases e.g. if they were informed about recordings by their general practitioners. In our study, we did not specify the extent to which “private” referred to non- or for-profit use, which limits a nuanced view on this aspect. Prior studies have demonstrated that intentions involving more commercial purposes, such as health insurance, are met with the greatest skepticism [4].

### Perspectives

Our findings have important implications for the design of data governance and AI policy in healthcare. The consistently higher willingness to share data with public institutions underscores the value of anchoring AI development within publicly governed infrastructures, such as secure hospital-based environments, and ensuring strong public oversight when private partners are involved. Differences in acceptance across data types further suggest that proportionate, tiered governance models may be warranted, with enhanced safeguards and consent procedures for more sensitive modalities such as genetic information and audio recordings. To address public reluctance to share data for AI purposes, there is a need to strengthen public willingness while respecting concerns and expectations. Such initiatives could promote the societal value of research, particularly its potential to improve population health, while ensuring that data sharing occurs under secure conditions. This includes transparent governance of data storage, clear control over data use, and effective communication of benefits and risks. Together, these measures can help align innovation with societal expectations and strengthen the trust necessary for sustainable and socially acceptable implementation of AI in healthcare.

## Conclusion

In conclusion, willingness to share health data for AI development is shaped by the perceived sensitivity of the data and the institutional context in which it is used. Greater acceptance of data sharing with public healthcare institutions, alongside substantial uncertainty regarding genetic and audio data, underscores the importance of trust, transparency, and clear communication about safeguards and purpose. As more data modalities are incorporated into AI-based solutions in healthcare, policies and governance frameworks that prioritize privacy protection, accountability, and public benefit, while actively engaging citizens and improving AI literacy, will be essential to ensure socially acceptable and sustainable implementation of AI in healthcare.

## Abbreviations

AI: artificial intelligence
GDPR: General Data Protection Regulation
HICD: Health in Central Denmark
LLM: large language model
OR: odds ratio
SD: standard deviation
T1D: type 1 diabetes
T2D: type 2 diabetes

## Acknowledgements

The authors are grateful to all participants of the Health in Central Denmark cohort and the steering committee, especially to Kasper Norman for his support with data management. The authors acknowledge the use of generative AI tools (ChatGPT by OpenAI) in the initial scoping of the study, and the writing process for revising the text (e.g. correct grammar, identify typos).

## Data availability

The Health in Central Denmark data is hosted on remote servers at Statistics Denmark. The project is managed by a steering committee at Steno Diabetes Center Aarhus, Aarhus University Hospital. The steering committee encourages interested researchers to use this resource. More information is available on the website: https://www.stenoaarhus.dk/research/resources/health-in-central-denmark/.

## Author contributions

J.R.S., A.A.I., and A.H. designed the questionnaire items on data sharing and designed the study. J.R.S. processed the data and conducted statistical analyses and visualizations with contributions from A.A.I. and A.H. All authors contributed to the initial draft of the manuscript. All authors contributed to data interpretation and provided feedback and contributions to the final manuscript. All authors agreed to the submission of the manuscript. All authors had full access to the data in the study.

## Supplementary materials

**Table S1.**
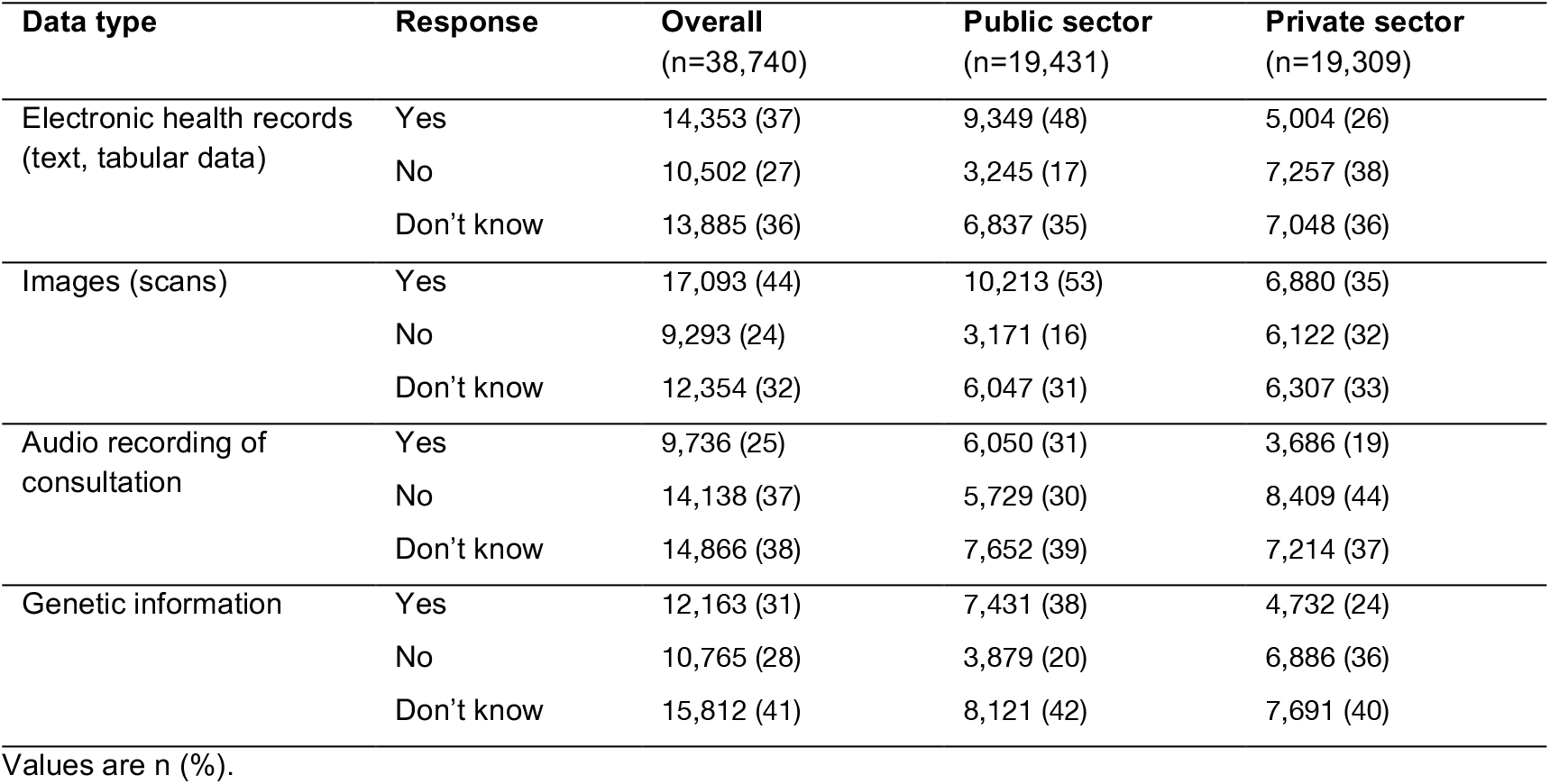
Distribution of data sharing attitudes by data type and sector (unweighted)

**Table S2.** Association (unweighted) between data recipient sector (public vs. private [reference]) and willingness to share data.

| Data type | Response | OR (95% CI) |
| --- | --- | --- |
| Electronic health records<br>(text, tabular data) | Yes | 1.93 (1.84, 2.02) |
|  | No | 0.46 (0.44, 0.49) |
|  | Don't know | 1 (reference) |
| Images (hospital scans) | Yes | 1.55 (1.48, 1.62) |
|  | No | 0.54 (0.51, 0.57) |
|  | Don't know | 1 (reference) |
| Audio recording of consultation | Yes | 1.55 (1.47, 1.63) |
|  | No | 0.64 (0.61, 0.67) |
|  | Don't know | 1 (reference) |
| Genetic information | Yes | 1.49 (1.42, 1.56) |
|  | No | 0.53 (0.51, 0.56) |
|  | Don't know | 1 (reference) |

## References

1. Raghupathi W, Raghupathi V. Big data analytics in healthcare: promise and potential. Health Information Science and Systems. 2014;2(1):3.

2. Pastorino R, De Vito C, Migliara G, Glocker K, Binenbaum I, Ricciardi W, et al. Benefits and challenges of Big Data in healthcare: an overview of the European initiatives. European Journal of Public Health. 2019;29(Supplement_3):23–7.

3. Topol EJ. As artificial intelligence goes multimodal, medical applications multiply. Science.381(6663):eadk6139.

4. Skovgaard L, Ekstrøm CT, Svendsen MN, Hoeyer K. Survey of attitudes in a Danish public towards reuse of health data. PLOS ONE. 2024;19(12):e0312558.

5. Olsen Q, Dyda A, Woods L, Lobo E, Eden R, Krahe MA, et al. Worldwide willingness to share health data high but privacy, consent and transparency paramount, a meta-analysis. npj Digital Medicine. 2025;8(1):540.

6. Middleton A, Milne R, Almarri MA, Anwer S, Atutornu J, Baranova EE, et al. Global Public Perceptions of Genomic Data Sharing: What Shapes the Willingness to Donate DNA and Health Data? The American Journal of Human Genetics. 2020;107(4):743–52.

7. Murdoch B. Privacy and artificial intelligence: challenges for protecting health information in a new era. BMC Medical Ethics. 2021;22(1):122.

8. Chakradeo K, Huynh I, Balaganeshan SB, Dollerup OL, Gade-Jørgensen H, Laupstad SK, et al. Navigating fairness aspects of clinical prediction models. BMC Medicine. 2025;23(1):567.

9. Amann J, Blasimme A, Vayena E, Frey D, Madai VI, the Precise Qc. Explainability for artificial intelligence in healthcare: a multidisciplinary perspective. BMC Medical Informatics and Decision Making. 2020;20(1):310.

10. Schaarup JFR, Aggarwal R, Dalsgaard E-M, Norman K, Dollerup OL, Ashrafian H, et al. Perception of artificial intelligence-based solutions in healthcare among people with and without diabetes: A cross-sectional survey from the health in Central Denmark cohort. Diabetes Epidemiology and Management. 2023;9:100114.

11. Fischer A, Delborg J. Forsker har fået lov at bruge millioner af danskeres sygehusjournaler - uden de ved det. DR. 2025.

12. Fischer A, Delborg J. Datatilsynet går ind i sag om brug af millioner af danskeres sygehusjournaler til AI. DR. 2025.

13. Bjerg L, Dalsgaard E-M, Norman K, Isaksen AA, Sandbæk A. Cohort profile: Health in Central Denmark (HICD) cohort - a register-based questionnaire survey on diabetes and related complications in the Central Denmark Region. BMJ Open. 2022;12(7):e060410.

14. Schaarup J, Isaksen AA, Hulman A. Peoples’ perception of artificial intelligence and chatbots: an online survey from Health in Central Denmark. Figshare. 2025; 10.6084/m9.figshare.27894180.

15. Isaksen AA, Sandbæk A, Bjerg L. Validation of Register-Based Diabetes Classifiers in Danish Data. Clin Epidemiol. 2023;15:569–81.

16. Canada Health I. 2024 Canadian Digital Health Survey. V3 ed: Borealis; 2025.

17. Mahmoud R, Moody AR, Foster M, Girdharry N, Sinn L, Zhang B, et al. Sharing De-identified Medical Images Electronically for Research: A Survey of Patients’ Opinion regarding Data Management. Canadian Association of Radiologists Journal. 2019;70(3):212–8.

18. Kim J, Kim H, Bell E, Bath T, Paul P, Pham A, et al. Patient Perspectives About Decisions to Share Medical Data and Biospecimens for Research. JAMA Network Open. 2019;2(8):e199550–e.

19. Chandrasekaran R, Moustakas E. Patient attitudes toward ambient artificial intelligence scribes in clinical care: insights from a cross-sectional study. Journal of the American Medical Informatics Association. 2026;33(2):263–72.

20. Leiserowitz G, Mansfield J, MacDonald S, Jost M. Patient Attitudes Toward Ambient Voice Technology: Preimplementation Patient Survey in an Academic Medical Center. JMIR Med Inform. 2025;13:e77901.

21. Aggarwal R, Farag S, Martin G, Ashrafian H, Darzi A. Patient Perceptions on Data Sharing and Applying Artificial Intelligence to Health Care Data: Cross-sectional Survey. J Med Internet Res. 2021;23(8):e26162.

22. Fesl S, Lang C, Schmitt J, Brückner S, Gilbert S, Deckert S, et al. Factors influencing patients’ willingness to share their digital health data for primary and secondary use: A theory-and evidence-based overview of reviews. Digital Health. 2025;11:20552076251340254.

23. Schaarup JR, Isaksen AA, Norman K, Bjerg L, Hulman A. Trust in large language model-based solutions in healthcare among people with and without diabetes: a cross-sectional survey from the Health in Central Denmark cohort. BMJ Digital Health & AI. 2025;1(1):e000090.

24. Isaksen AA, Schaarup JR, Bjerg L, Hulman A. Changes in public perception of artificial intelligence in healthcare after exposure to ChatGPT. npj Digital Medicine. 2025;8(1):795.

25. Kuo R, O’Hanlon C, Smith J, Hill R, Papiez BW, Collins GS, et al. Public perceptions of health data sharing for artificial intelligence research: a qualitative focus group study in the UK. BMJ Digital Health & AI. 2026;2(1):e000239.

26. Cascini F, Pantovic A, Al-Ajlouni YA, Puleo V, De Maio L, Ricciardi W. Health data sharing attitudes towards primary and secondary use of data: a systematic review. eClinicalMedicine. 2024;71.

27. The European Parliament and the Council of the European Union. General Data Protection Regulation. 2016.

28. Ganna A, Carracedo A, Christiansen CF, Di Angelantonio E, Dykstra PA, Dzhambov AM, et al. The European Health Data Space can be a boost for research beyond borders. Nature Medicine. 2024;30(11):3053–6.

29. Busch F, Kather JN, Johner C, Moser M, Truhn D, Adams LC, et al. Navigating the European Union Artificial Intelligence Act for Healthcare. npj Digital Medicine. 2024;7(1):210.

